# Parental Socio-economic Class and Obesity in Hispanic and non-Hispanic Individuals with First Episode Psychosis

**DOI:** 10.1101/2025.09.22.25336105

**Authors:** Santiago Vega-Ramos, Santiago Alvarez-Lesmes, Krisha Arora-Guevara, Kelly García-Bohórquez, Mauricio Tohen, Todd Lencz, Anil K. Malhotra, Juan A. Gallego

## Abstract

**Introduction:** Obesity is a significant public health issue in the United States (U.S.), with rates of obesity increasing for the past few years, particularly in Hispanics. Individuals with schizophrenia have additional risks for obesity due to the metabolic burden associated with antipsychotic (AP) medications, limited physical activity due to negative symptoms, and poor eating habits. There are no published studies that have compared obesity rates between Hispanics and non-Hispanics with first episode psychosis (FEP). We aimed to explore the relationship between ethnicity and obesity in FEP patients prior to the initiation of AP treatment, to eliminate the potential effects of AP treatment on weight.

**Methods:** Baseline data from 145 individuals with FEP enrolled in a FEP research study was used. Demographic, education, occupation and Body Mass Index (BMI) data for FEP participants was stratified by ethnicity (Hispanic vs. non-Hispanic) and BMI (< 25 or ≥ 25). Variables with a p-value < 0.1 were entered into a multivariate linear regression model using a manual backwards elimination approach, using BMI as a continuous measure as the outcome and ethnicity as our predictor of interest.

**Results:** Twenty-four Hispanics, and 121 non-Hispanics were included. Baseline characteristics were not significantly different between ethnicity groups, except for BMI which was significantly higher in Hispanics (mean = 25.3, SD = 6.0, p = 0.037) than non-Hispanics (mean = 23.0, SD = 4.5). Data stratified by BMI showed that age, ethnicity, mother’s occupation, and socioeconomic status (SES) were associated with BMI. Multivariate linear regression showed that Hispanic ethnicity (B = 3.04, SE = 1.32, p = 0.024) and age (B = 0.36, SE = 0.09, p < 0.001) were statistically significantly associated with higher BMI scores while adjusting for sex (B = - 0.67, SE = 1.04, p = 0.521).

**Discussion:** Hispanic individuals with FEP present with higher BMI scores compared to non-Hispanics, even before the initiation of antipsychotic treatment. Therefore, in addition to exercise and healthy eating habits, the use of AP medications with less metabolic burden is advisable along with using the lowest effective dose.

## Introduction

Obesity, one of the most compelling public health concerns in the United States (U.S.), has been on the rise over the last few decades. According to the U.S. Centers for Disease Control and Prevention (CDC), the most recent report on obesity in adults revealed a prevalence of 40.3% (Emmerich et al., 2024). Obesity is a chronic illness associated with serious adverse outcomes such as increased rates of cardiovascular disease (Cercato & Fonseca, 2023), diabetes (Banday et al., 2022), and hyperlipidemia (Chooi et al., 2022). Unfortunately, due to those health care disparities and sociocultural factors, certain ethnic groups, such as the minority Hispanic population in the U.S., exhibit higher rates of obesity than others (Alemán et al., 2023; Iruretagoyena et al., 2019).

Compared to non-Hispanics, Hispanics generally consume diets higher in added sugars, saturated fats, and sodium, while lower in fruits, vegetables, and fiber. These dietary patterns are more pronounced among U.S.-born Hispanics, who tend to adopt Westernized food practices, a process that has been associated with increased rates of obesity (Akresh, 2007; Da Costa, 2023), diabetes (Koh, 2023), and cardiovascular disease (Chen et al., 2022; Isasi et al., 2015; Maldonado et al., 2021). Additionally, Hispanic adults in the U.S. report lower physical activity levels than non-Hispanic adults (CDC, 2022). Barriers include lack of time, limited access to recreational spaces, economic stress, cultural responsibilities, and perceptions that healthy foods are costly or unappealing (Ghaddar et al., 2010; Heredia et al., 2022).

Obesity is frequently seen in individuals with schizophrenia, largely due to the metabolic burden linked to antipsychotic (AP) medications, poor dietary patterns, and a sedentary lifestyle driven by negative symptoms. These negative symptoms — such as avolition, anhedonia, and social withdrawal — contribute to diminished motivation and reduced engagement in physical activities, which further exacerbate weight gain. Several studies have found a higher prevalence of obesity in people diagnosed with schizophrenia when compared to healthy controls (Annamalai et al., 2017; Iruretagoyena et al., 2019; Li et al., 2023; Smith et al., 2020; Tian et al., 2021). This is true not only for patient with chronic schizophrenia but also in patients with first episode psychosis (FEP) (Iruretagoyena et al., 2019; Li et al., 2023; Smith et al., 2020; Tian et al., 2021). Individuals experiencing FEP commonly exhibit substantial increases in body weight and metabolic abnormalities, which can be intensified by AP medications such as clozapine and olanzapine (Annamalai et al., 2017; Iruretagoyena et al., 2019; Smith et al., 2020; Tian et al., 2021).

To date, only one study has reported on metabolic disturbances in Hispanic FEP individuals (Iruretagoyena et al., 2019), but this study took place in Chile and did not have a non-Hispanic control group. We did not find any studies directly comparing obesity rates between Hispanic and non-Hispanic FEP individuals living in the U.S, and prior to the initiation of AP treatment. To fill this gap, we conducted a post-hoc analysis of baseline data of FEP individuals that participated in a 12-week long trial. By focusing on FEP population and prior to the start of AP treatment for the clinical trial, we aim to determine the impact of ethnicity on obesity, while minimizing the impact of AP treatment on weight and body mass index (BMI). We hypothesize that Hispanic FEP patients will have higher BMI compared to non-Hispanic FEP patients.

## Methods

For this post-hoc analysis, we used baseline data collected from 145 individuals with FEP that participated in the Striatal Connectivity and Clinical Outcome in Psychosis study (R01MH108654, PI: Malhotra) (ClinicalTrials.gov ID: NCT02822092) at Zucker Hillside Hospital in NY, between 2016 and 2021.

### Inclusion and Exclusion Criteria

Patients were included in the study if they: (1) fulfilled DSM-5 criteria for schizophrenia, schizophreniform disorder, schizoaffective disorder, psychosis not otherwise specified, brief psychotic disorder, bipolar I with psychotic features (acute manic or mixed episode), major depressive disorder with psychotic features or substance induced psychotic disorders; (2) did not meet DSM-5 criteria for a current psychotic disorder due to a general medical condition, delusional disorder, shared psychotic disorder, or a mood disorder without psychotic features; (3) had current positive symptoms rated ≥4 (moderate) on one or more of these BPRS items: conceptual disorganization, grandiosity, hallucinatory behavior, unusual thought content; (4) were in an early phase of illness as defined by having taken AP medications for a cumulative lifetime period of 4 weeks or less; (5) were 15-40 years old; (6) were willing and capable of providing informed consent; and (7) were fluent in English.

Subjects were excluded from participation if they (1) had a serious neurological or endocrine disorder or any medical condition or treatment known to affect the brain; (2) had any medical condition which required treatment with a medication with psychotropic effects; (3) displayed significant risk of suicidal or homicidal behavior; (4) experienced cognitive or language limitations, or any other factor that would preclude subjects providing informed consent; (5) had medical contraindications to treatment with risperidone or aripiprazole, (6) had a lack of response to a prior adequate trial of risperidone or aripiprazole, or (7) experienced Magnetic resonance imaging (MRI) contraindications.

#### Assessments

Psychopathology assessments included the Brief Psychiatric Rating Scale-anchored version (BPRS-A) (Ventura et al., 1993), the Schedule for Assessment of Negative Symptoms (SANS) (Andreasen, 1989), and the Global Assessment Scale (GAS) (Endicott et al., 1976). SES was assessed using the Hollingshead Two-Factor Index, which classifies individuals into five categories: lower, lower middle, middle, upper middle, and upper class. A composite score was calculated based on the education and occupation of each participant and their parents, providing a standardized measure of social status.

### Statistical Analysis

All statistical analyses were conducted using R (version 4.4.1) within the RStudio environment. Analyses were performed using the following R packages: dplyr for data manipulation, tableone for descriptive statistics, broom for tidying model outputs, and emmeans for post-hoc comparisons.

Data were initially stratified by ethnicity (Hispanic vs. non-Hispanic). Baseline characteristics—including demographics, education, occupation, socioeconomic status (SES), BMI, and parental education, parental occupation, and parental SES—were compared using independent t-tests for normally distributed continuous variables and Wilcoxon rank-sum tests for non-normally distributed variables. Chi-square tests or Fisher’s exact tests (for small cell counts) were used to compare categorical variables. Bivariate analyses for the same variables were then performed comparing those participants with BMI< 25 with those ≥ 25. A BMI of 25 or higher was selected mirroring the CDC classification which categorizes as overweight or obese those individuals with a BMI of 25 or more.

Multivariate modeling: We modeled BMI as a continuous outcome, treating ethnicity as the primary predictor of interest. Age and sex were included a priori. We then considered additional covariates based on prior literature and our bivariable screening using a p-value cutoff of < 0.10. From there, we used a manual backward elimination approach to arrive at a parsimonious model, removing variables one at a time using the largest p-value as the criteria for removal, except for sex and age. After arriving to our parsimonious model, we tested for potential confounders and left those variables in the model if they modified the B-estimate for our main predictor of interest (ethnicity) by >20%. To ensure valid comparison between models with and without the potential confounder, we restricted both models to the same subset of participants with complete data for all variables (n=91).

## Results

### Sample Characteristics

A total of 145 individuals with FEP were included in the analysis. Baseline demographic and clinical characteristics—including sex, age, marital status, education, occupation, SES, BPRS score, GAS score, and THC use—did not show any statistically significant difference between Hispanic and non-Hispanic groups. Importantly, English was the primary language for both Hispanics and non-Hispanics. On the other hand, BMI was statistically significantly higher among Hispanic participants (mean = 25.34, SD = 5.95) compared to non-Hispanics (mean = 22.95, SD = 4.46; p = 0.037) (Table 1).

**Table 1.**
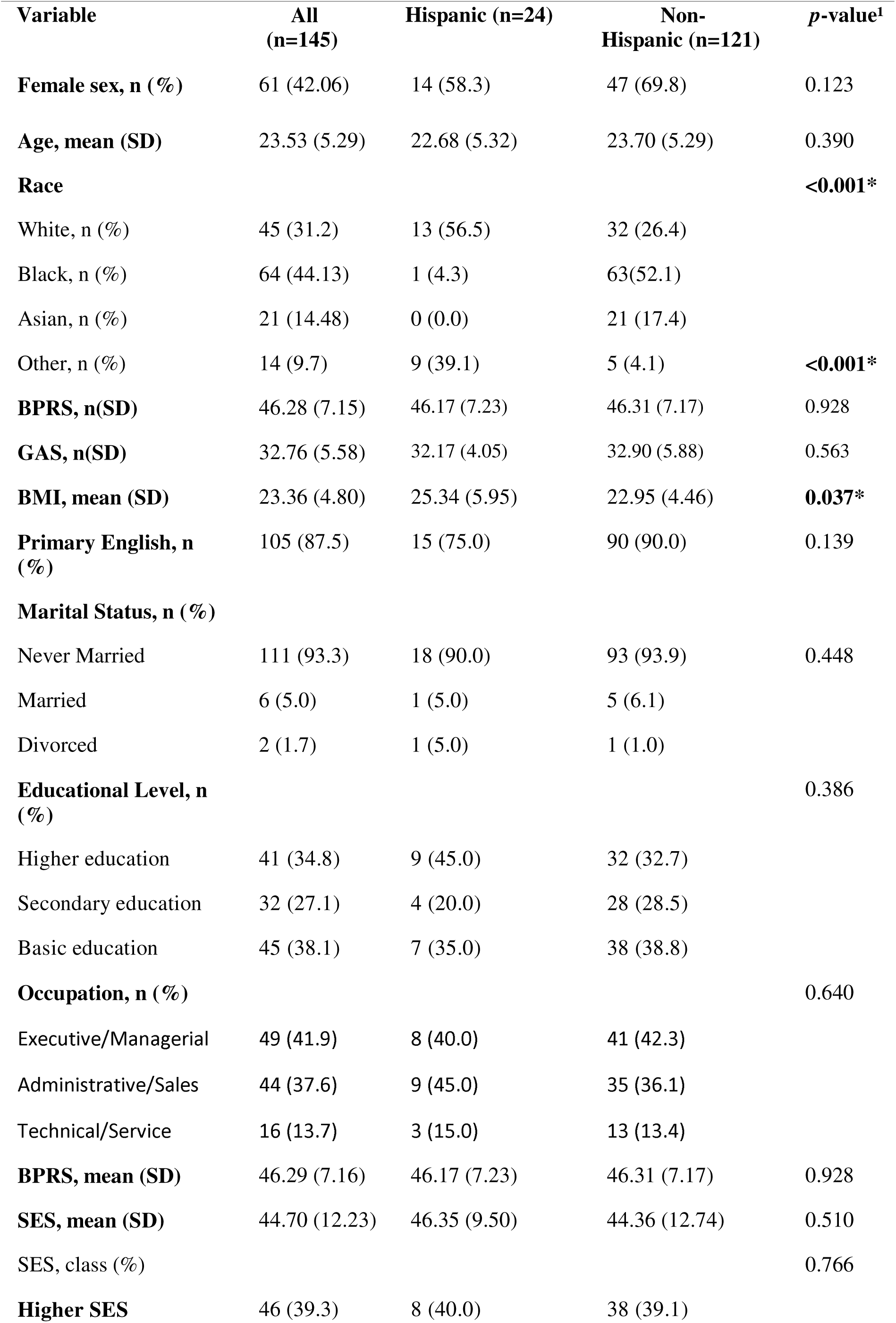
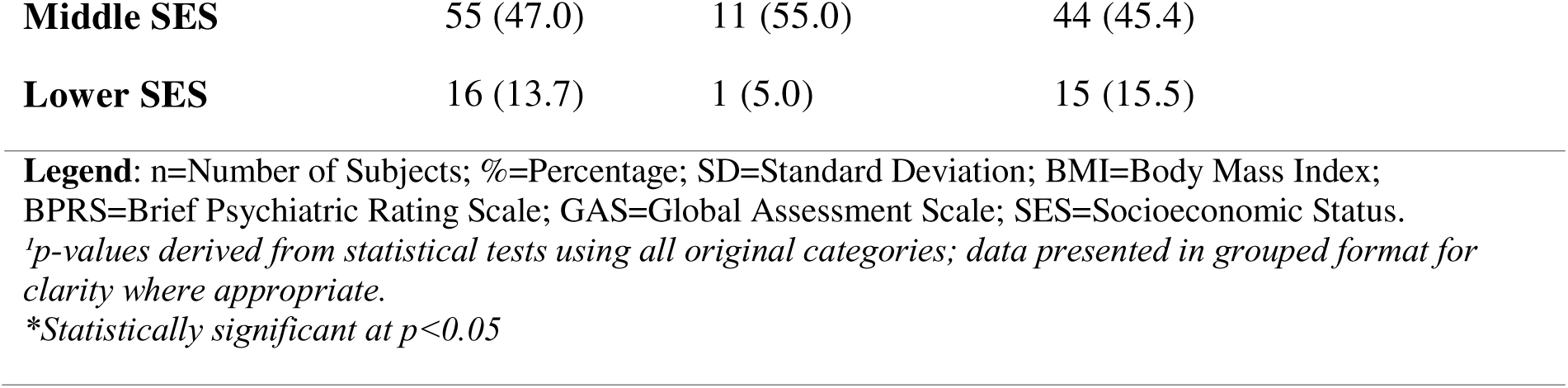
Baseline Characteristics by Ethnicity Group.

| Variable | All<br>(n=145) | Hispanic (n=24) | Non-Hispanic (n=121) | *p*-value<sup>1</sup> |
| --- | --- | --- | --- | --- |
| **Female sex, n (%)** | 61 (42.06) | 14 (58.3) | 47 (69.8) | 0.123 |
| **Age, mean (SD)** | 23.53 (5.29) | 22.68 (5.32) | 23.70 (5.29) | 0.390 |
| **Race** |  |  |  | **<0.001\*** |
| White, n (%) | 45 (31.2) | 13 (56.5) | 32 (26.4) |  |
| Black, n (%) | 64 (44.13) | 1 (4.3) | 63(52.1) |  |
| Asian, n (%) | 21 (14.48) | 0 (0.0) | 21 (17.4) |  |
| Other, n (%) | 14 (9.7) | 9 (39.1) | 5 (4.1) | **<0.001\*** |
| **BPRS, n(SD)** | 46.28 (7.15) | 46.17 (7.23) | 46.31 (7.17) | 0.928 |
| **GAS, n(SD)** | 32.76 (5.58) | 32.17 (4.05) | 32.90 (5.88) | 0.563 |
| **BMI, mean (SD)** | 23.36 (4.80) | 25.34 (5.95) | 22.95 (4.46) | **0.037\*** |
| **Primary English, n (%)** | 105 (87.5) | 15 (75.0) | 90 (90.0) | 0.139 |
| **Marital Status, n (%)** |  |  |  |  |
| Never Married | 111 (93.3) | 18 (90.0) | 93 (93.9) | 0.448 |
| Married | 6 (5.0) | 1 (5.0) | 5 (6.1) |  |
| Divorced | 2 (1.7) | 1 (5.0) | 1 (1.0) |  |
| **Educational Level, n (%)** |  |  |  | 0.386 |
| Higher education | 41 (34.8) | 9 (45.0) | 32 (32.7) |  |
| Secondary education | 32 (27.1) | 4 (20.0) | 28 (28.5) |  |
| Basic education | 45 (38.1) | 7 (35.0) | 38 (38.8) |  |
| **Occupation, n (%)** |  |  |  | 0.640 |
| Executive/Managerial | 49 (41.9) | 8 (40.0) | 41 (42.3) |  |
| Administrative/Sales | 44 (37.6) | 9 (45.0) | 35 (36.1) |  |
| Technical/Service | 16 (13.7) | 3 (15.0) | 13 (13.4) |  |
| **BPRS, mean (SD)** | 46.29 (7.16) | 46.17 (7.23) | 46.31 (7.17) | 0.928 |
| **SES, mean (SD)** | 44.70 (12.23) | 46.35 (9.50) | 44.36 (12.74) | 0.510 |
| SES, class (%) |  |  |  | 0.766 |
| **Higher SES** | 46 (39.3) | 8 (40.0) | 38 (39.1) |  |

|  |  |  |  |
| --- | --- | --- | --- |
| **Middle SES** | 55 (47.0) | 11 (55.0) | 44 (45.4) |
| **Lower SES** | 16 (13.7) | 1 (5.0) | 15 (15.5) |

**Legend:** n=Number of Subjects; %=Percentage; SD=Standard Deviation; BMI=Body Mass Index; BPRS=Brief Psychiatric Rating Scale; GAS=Global Assessment Scale; SES=Socioeconomic Status. <sup>1</sup>*p-values derived from statistical tests using all original categories; data presented in grouped format for clarity where appropriate.* *\*Statistically significant at $p<0.05$*

### Parental Education and Occupation

There were significant differences in the father’s occupation between Hispanic and non-Hispanic FEP individuals (p=0.037). For example, 70.5% (n = 12; p = 0.037) of Hispanic fathers were employed in skilled manual, semi-skilled, or unskilled labor roles, compared to 37.9% (n = 33) of non-Hispanic fathers. None of the Hispanic fathers were classified as high-level executives or business managers, whereas 38.8% (n = 33) of non-Hispanic fathers held such positions.

There were also significant differences in mother’s occupation between groups (p<0.001). A 52.7% (n = 10; p < 0.001) of Hispanic mothers were in skilled manual, semi-skilled, or unskilled roles, compared to 26.7% (n = 24) of non-Hispanic mothers and only 5.3% (n = 1) of Hispanic mothers were business managers, compared to 37.8% (n = 34) of non-Hispanic mothers (Table 2).

**Table 2.**
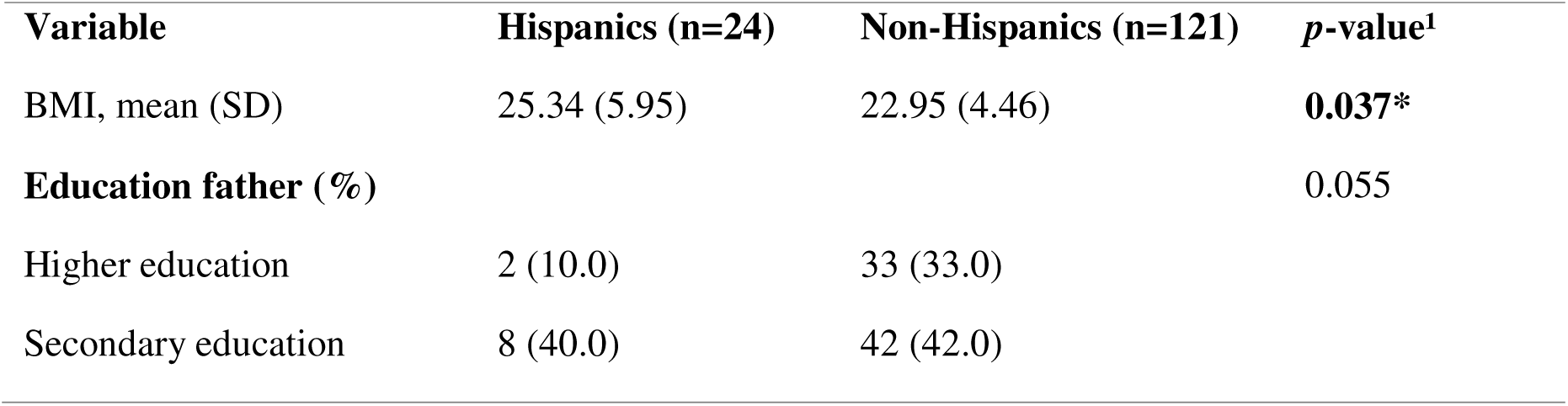
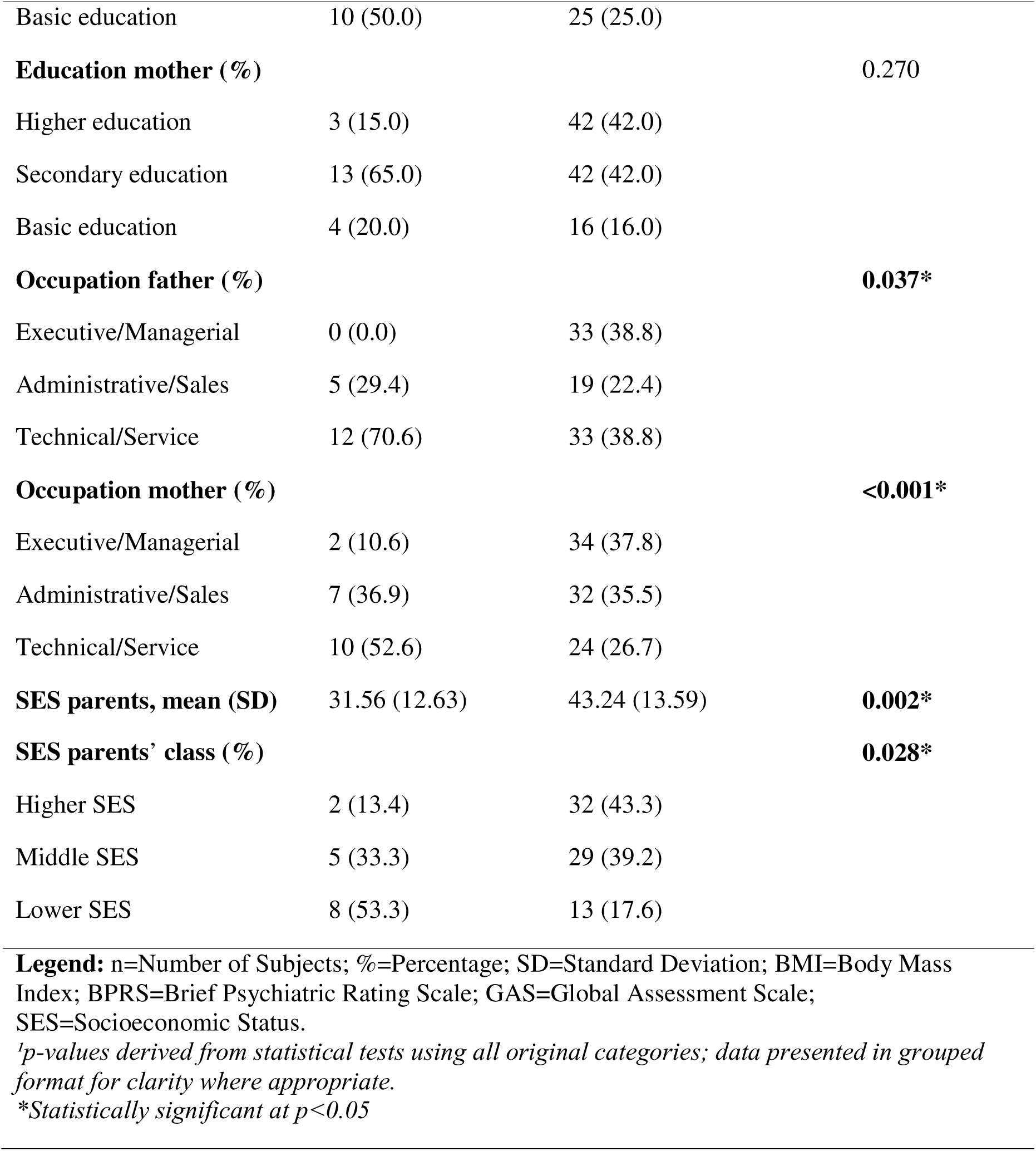
Baseline Characteristics by Ethnicity group – Parents.

| **Variable** | **Hispanics (n=24)** | **Non-Hispanics (n=121)** | ***p*-value<sup>1</sup>** |
| --- | --- | --- | --- |
| BMI, mean (SD) | 25.34 (5.95) | 22.95 (4.46) | **0.037\*** |
| **Education father (%)** |  |  | 0.055 |
| Higher education | 2 (10.0) | 33 (33.0) |  |
| Secondary education | 8 (40.0) | 42 (42.0) |  |
| Basic education | 10 (50.0) | 25 (25.0) |  |
| **Education mother (%)** |  |  | 0.270 |
| Higher education | 3 (15.0) | 42 (42.0) |  |
| Secondary education | 13 (65.0) | 42 (42.0) |  |
| Basic education | 4 (20.0) | 16 (16.0) |  |
| **Occupation father (%)** |  |  | **0.037\*** |
| Executive/Managerial | 0 (0.0) | 33 (38.8) |  |
| Administrative/Sales | 5 (29.4) | 19 (22.4) |  |
| Technical/Service | 12 (70.6) | 33 (38.8) |  |
| **Occupation mother (%)** |  |  | **<0.001\*** |
| Executive/Managerial | 2 (10.6) | 34 (37.8) |  |
| Administrative/Sales | 7 (36.9) | 32 (35.5) |  |
| Technical/Service | 10 (52.6) | 24 (26.7) |  |
| **SES parents, mean (SD)** | 31.56 (12.63) | 43.24 (13.59) | **0.002\*** |
| **SES parents' class (%)** |  |  | **0.028\*** |
| Higher SES | 2 (13.4) | 32 (43.3) |  |
| Middle SES | 5 (33.3) | 29 (39.2) |  |
| Lower SES | 8 (53.3) | 13 (17.6) |  |
| **Legend:** n=Number of Subjects; %=Percentage; SD=Standard Deviation; BMI=Body Mass Index; BPRS=Brief Psychiatric Rating Scale; GAS=Global Assessment Scale; SES=Socioeconomic Status.<br>*<sup>1</sup>p-values derived from statistical tests using all original categories; data presented in grouped format for clarity where appropriate.*<br>*\*Statistically significant at p<0.05* |  |  |  |

### Socioeconomic Status (SES)

Based on the Hollingshead Two-Factor Index, 86.6% (n = 13; p = 0.028) of Hispanic parents were classified in SES Classes II–III (Low to Middle), while only 13.4% (n = 2) were in Classes IV–V (High to Very High). In comparison, 55.4% (n = 31) of non-Hispanic parents were in Classes II–III, and 43.3% (n = 32; p=0.028)) were in Classes IV–V.

### Stratified Analysis by BMI

When comparing participants with BMI < 25 vs. ≥ 25, those with BMI ≥ 25 were older on average (mean 25.96 years, SD 6.82) than those with BMI < 25 (mean 22.85 years, SD 4.52; p = 0.004). A higher proportion of individuals with BMI ≥ 25 were Hispanic (12/21, 57.1%; p = 0.005), compared to non-Hispanic (24/102; 23.5% p = 0.005). Participants with BMI ≥ 25 were more likely to have mothers in semi-skilled or skilled manual jobs (53.1%; p = 0.001), while those with BMI < 25 more commonly had mothers in administrative roles (29.2%; p = 0.001) or business management positions (40.3%; p = 0.001). Finally, participants with BMI ≥ 25 had a lower average parental SES score (mean=36.08, SD 14.62) compared with those with BMI < 25 (mean=43.58, SD 12.84; p = 0.018). (Table 3).

**Table 3.** Stratified by BMI ≥ 25 and < 25.

| **Table 3. Stratified by BMI ≥ 25 and < 25** |  |  |  |
| --- | --- | --- | --- |
| **Variable** | **BMI ≥ 25 (n=36)** | **BMI < 25 (n=87)** | **p-value<sup>1</sup>** |
| **Age, mean (SD)** | 25.96 (6.82) | 22.85 (4.52) | **0.004\*** |
| **Ethnicity** |  |  | **0.005\*** |
| Hispanic | 12/21 (57.1.) | 9/21 (42.9.3) |  |
| Non-Hispanic | 24/102 (23.5) | 78/102 (76.5) |  |
| **Female sex, n (%)** | 15 (41.7) | 36 (41.4) | 1.000 |
| **Race (%)** |  |  | 0.527 |
| Asian or Pacific Islander | 3 (8.6) | 13 (14.9) |  |
| Black | 14 (40.0) | 41 (47.1) |  |
| White | 13 (37.1) | 25 (28.7) |  |
| Other | 5 (14.3) | 8 (9.2) |  |
| **BPRS, mean (SD)** | 44.47 (6.31) | 46.58 (7.15) | 0.129 |
| **GAS, mean (SD)** | 32.34 (5.40) | 32.95 (5.65) | 0.588 |
| **Marital status (%)** |  |  | 0.101 |
| Single | 29 (85.3) | 77 (96.2) |  |
| Married/Divorced | 5 (14.7) | 3 (3.8) |  |
| **Occupation (%)** |  |  | 0.277 |
| Executive/Managerial | 18 (54.6) | 29 (36.7) |  |
| Administrative/Sales | 10 (30.4) | 32 (40.5) |  |
| Technical/Service | 5 (15.1) | 18 (22.8) |  |
| Unemployed | 0 (0.0) | 4 (5.1) |  |
| **Education (%)** |  |  | 0.111 |
| Higher education | 10 (30.3) | 30 (37.5) |  |
| Secondary education | 5 (15.1) | 26 (32.5) |  |
| Basic education | 18 (54.5) | 24 (30.0) |  |
| **SES, mean (SD)** | 46.76 (11.09) | 43.77 (12.59) | 0.239 |
| **English Primary Language (%)** | 28 (82.4) | 72 (90.0) | 0.409 |
| **Education father (%)** |  |  | 0.141 |
| Higher education | 9 (26.5) | 24 (30.0) |  |
| Secondary education | 12 (35.3) | 36 (45.0) |  |
| Basic education | 11 (32.3) | 20 (25.0) |  |
| **Education mother (%)** |  |  | 0.191 |
| Higher education | 8 (24.2) | 36 (44.4) |  |
| Secondary education | 16 (48.5) | 34 (42.0) |  |
| Basic education | 9 (27.3) | 11 (13.6) |  |
| **Occupation father (%)** |  |  | 0.920 |
| Executive/Managerial | 8 (27.6) | 23 (33.8) |  |
| Administrative/Sales | 6 (20.7) | 17 (25.0) |  |
| Technical/Service | 15 (51.7) | 28 (41.2) |  |
| **Occupation mother (%)** |  |  | **0.001\*** |
| Executive/Managerial | 4 (12.5) | 29 (40.3) |  |
| Administrative/Sales | 11 (34.4) | 28 (38.9) |  |
| Technical/Service | 17 (53.1) | 15 (20.8) |  |
| **SES parents (mean (SD))** | 36.08 (14.62) | 43.58 (12.84) | **0.018\*** |

**Legend:** n=Number of Subjects; %=Percentage; SD=Standard Deviation; BMI=Body Mass Index; BPRS=Brief Psychiatric Rating Scale; GAS=Global Assessment Scale; SES=Socioeconomic Status.
*<sup>1</sup>p-values derived from statistical tests using all original categories; data presented in grouped format for clarity where appropriate.*
*\*Statistically significant at $p < 0.05$*

### Multivariable Linear Regression Analysis

The initial model included ethnicity, age, sex, and parental SES. We did not include maternal occupation in the initial model because it is a component of the parental SES index to avoid multicollinearity. Using a manual backward elimination approach, the final parsimonious model showed that being non-Hispanic was statistically significantly associated with a 3.04 kg/m² lower BMI, compared to Hispanic participants (β = −3.04, SE = 1.32, p = 0.024) after adjusting for age and sex. Age was positively associated with BMI, with each additional year of age associated with a 0.36 kg/m² increase in BMI (β = 0.36, SE = 0.09, p < 0.001). Sex was not significantly associated with BMI (β = −0.67, SE = 1.04, p = 0.521), but was left in the model to account for the potential effects of sex in the outcome. The final model explained approximately 18% of the variance in BMI (N = 91; R² = 0.18; F(3, 87) = 6.53; p < 0.001).

To assess whether parental SES confounded the ethnicity–BMI association, we fit the same model with the addition of parental SES on the same subset of participants with complete data (N = 91). In this SES-adjusted model, the ethnicity coefficient was attenuated from β = - 3.04 to β = −2.37 (SE = 1.40; p = 0.095), representing a 22% decrease in magnitude and a loss of statistical significance. This exceeded our prespecified threshold for confounding (20% change in β coefficient), so parental SES was retained in the model. In this model, age remained positively associated with BMI (β = 0.33, SE = 0.09, p < 0.001), sex was not associated with BMI (β = −0.48, SE = 1.04, p = 0.644), and parental SES did not show an independent association with BMI in the presence of the other variables (β = −0.05, SE = 0.04, p = 0.161). Model fit for the SES-adjusted model was R² = 0.20 (N = 91; F(4, 86) = 5.37; p < 0.001), indicating that 20% of the variance in BMI was explained by the model. (Table 4).

**Table 4.** Models of linear regression excluding and including parental SES.

| Variable | Coefficient (B) | SE | 95% CI | t | p-value |
| --- | --- | --- | --- | --- | --- |
| *Model 1: Without adjustment for parental SES* |  |  |  |  |  |
| Intercept | 17.93 | 2.40 | [13.16, 22.69] | 7.46 | <0.001 |
| Non-Hispanic ethnicity | -3.04 | 1.32 | [-5.67, -0.41] | -2.29 | 0.024 |
| Age | 0.36 | 0.09 | [0.17, 0.54] | 3.83 | <0.001 |
| Sex | -0.67 | 1.04 | [-2.73, 1.39] | -0.65 | 0.521 |
| *Model 2: With adjustment for parental SES* |  |  |  |  |  |
| Intercept | 20.00 | 2.80 | [14.43, 25.57] | 7.14 | <0.001 |
| Non-Hispanic ethnicity | -2.37 | 1.40 | [-5.15, 0.41] | -1.69 | 0.095 |
| Age | 0.33 | 0.09 | [0.15, 0.52] | 3.57 | <0.001 |
| Sex | -0.48 | 1.04 | [-2.55, 1.59] | -0.46 | 0.644 |
| Parental SES | -0.05 | 0.04 | [-0.13, 0.02] | -1.42 | 0.161 |

$n=91$ ; Model 1: $R^2=0.18$ , $F(3,87)=6.53$ , $p<0.001$ ; Model 2: $R^2=0.20$ , $F(4,86)=5.37$ , $p<0.001$
**Legend:** SE=Standard Error; CI=Confidence Interval; SES=Socioeconomic Status; BMI=Body Mass Index. \*Statistically significant at $p<0.05$

## Discussion

To our knowledge, this is the first report comparing obesity rates between Hispanic and non-Hispanic FEP individuals living in the U.S. As hypothesized, we initially found significantly elevated BMI values in Hispanic FEP participants compared to their non-Hispanic counterparts. However, this finding became non-significant in our multivariate regression model once parental socio-economic class was added to the model.

Our finding highlights the critical role of social determinants of health in shaping metabolic outcomes in this population, rather than ethnicity per se. Specifically, Hispanic participants’ parents, particularly mothers, had lower levels of education and SES compared to non-Hispanic parents. Although our sample likely consists of second-generation Hispanics (as their primary language was English), their parents are likely first-generation immigrants who faced significant socioeconomic challenges, as evidenced by the marked disparities in social determinants of health observed in our bivariate analysis.

Our finding that parental SES modified the effects of ethnicity on BMI has been reported in general population studies but rarely examined in FEP populations. These findings align with broader evidence demonstrating that the Hispanic community bears a disproportionately high burden of obesity, driven largely by socioeconomic disparities and structural barriers to healthy living (Akresh, 2007; Alemán et al., 2023). The observed differences in BMI likely stem not from ethnicity itself, but from broader social determinants of health, such as intergenerational education, economic inequality, and systemic access barriers. Several studies have shown that diet and exercise habits change substantially after migration to the U.S., contributing to increased cardiovascular risk (Goel et al., 2004; Maldonado et al., 2021), with acculturation linked to lower diet quality, particularly among U.S.-born Hispanics or those who have lived longer in the U.S. (Chen et al., 2022; Isasi et al., 2015). This is consistent with literature demonstrating how lower SES, limited time and space for physical activity, and cost-related barriers to healthy food options are more common in Hispanic communities (Ghaddar et al., 2010; Heredia et al., 2022; Vatavuk-Serrati et al., 2023). Our study suggests that elevated BMI in Hispanic patients with FEP may reflect these broader lifestyle patterns and systemic inequities rather than inherent biological ethnic differences.

This fills an important gap in the literature, as most existing studies either do not stratify by ethnicity or only include patients with chronic schizophrenia, obscuring key differences (George et al., 2014). Strengths of our study include the inclusion of FEP participants with minimal or no exposure to AP medications, a large sample size for a single site FEP study, a comprehensive characterization of demographic and clinical characteristics at baseline, and the use of multivariate model. However, this study also has some limitations. While we adjust for several sociodemographic variables, unmeasured factors such as acculturation level, food insecurity, and physical activity habits were not collected and could not be included in our analyses and could further explain BMI differences.

Given these insights, targeted recommendations for clinical practice and public health interventions are warranted. Efforts to address obesity risk in Hispanics with FEP should go beyond individual-level behavior change and incorporate culturally informed, community-based strategies. Interventions should address barriers such as affordability, access to fresh food, and safe spaces for physical activity. Programs tailored to Hispanic communities that offer nutrition education, culturally relevant healthy recipes, and group-based physical activity may be especially effective. Additionally, early psychoeducation on diet and exercise during the initial phase of psychosis could mitigate long-term metabolic burden.

Similarly, structural interventions aimed at improving parental socioeconomic opportunities, such as access to higher education, job training, and fair employment practices, may have downstream effects on the metabolic health of younger generations. Such upstream approaches could complement individual-level interventions to create more sustainable improvements in health outcomes.

Future research should explore the intersection of ethnicity, acculturation, and social determinants of health in FEP individuals, not only in Hispanic individuals but also in other disadvantaged immigrant groups, given that it is likely that a similar effect may be seen in those groups. Studies should incorporate validated dietary and physical activity assessments adapted for Spanish-speaking populations to enhance the accuracy and cultural relevance of findings. Additionally, examining the relative contributions of parental versus individual SES across different developmental periods could provide insights into critical windows for intervention.

In conclusion, this study highlights the impact of social and economic factors in rates of obesity among individuals with FEP. While Hispanic ethnicity was initially associated with higher BMI, this effect was attenuated after adjusting for parental SES, underscoring the importance of addressing health disparities through a lens of social determinants rather than race or ethnicity alone. This finding may extend beyond Hispanics to other ethnic groups with lower SES. Culturally tailored, multilevel interventions are needed to reduce the metabolic burden among patients from disadvantaged backgrounds with early psychosis and to ensure equitable health outcomes.

## Data Availability

All data produced in the present study are available upon reasonable request to the authors.

## Acknowledgements

None

## References

Alemán, J. O., Almandoz, J. P., Frias, J. P., & Galindo, R. J. (2023). Obesity among Latinx people in the United States: A review. Obesity (Silver Spring), 31(2), 329–337. 10.1002/oby.23638

Akresh, I. R. (2007). Dietary assimilation and health among Hispanic immigrants to the United States. Journal of Health and Social Behavior, 48(4), 404–417. 10.1177/002214650704800405

Andreasen, N. C. (1989). The Scale for the Assessment of Negative Symptoms (SANS): Conceptual and theoretical foundations. The British Journal of Psychiatry, 155(S7), 49–52. 10.1192/S0007125000291496

Annamalai, A., Kosir, U., & Tek, C. (2017). Prevalence of obesity and diabetes in patients with schizophrenia. World Journal of Diabetes, 8(8), 390–396. 10.4239/wjd.v8.i8.390

Banday, M. Z., Sameer, A. S., & Nissar, S. (2022). Pathophysiology of obesity: Why is it so serious? Diabetes & Metabolic Syndrome: Clinical Research & Reviews, 16(1), 102371. 10.1016/j.dsx.2021.102371

CDC. (2022). Physical activity among adults: United States, 2020. https://www.cdc.gov/nchs/products/databriefs/db443.htm

Cercato, C., & Fonseca, F. A. (2023). Cardiovascular risk and obesity. Frontiers in Endocrinology, 14, 1130320. 10.3389/fendo.2023.1130320

Chen, Y. Y., Chen, G. C., Abittan, N., Xing, J., Mossavar Rahmani, Y., Sotres Alvarez, D., Mattei, J., Daviglus, M., Isasi, C. R., Hu, F. B., Kaplan, R., & Qi, Q. (2022). Healthy dietary patterns and risk of cardiovascular disease in US Hispanics/Latinos: The Hispanic Community Health Study/Study of Latinos (HCHS/SOL). American Journal of Clinical Nutrition, 116(4), 920–927. 10.1093/ajcn/nqac199

Chooi, Y. C., Ding, C., Chan, Z., Choo, J., Sadananthan, S. A., Michael, N., Lee, Y., Velan, S. S., & Magkos, F. (2022). Moderate weight loss improves body composition and metabolic function in metabolically unhealthy lean subjects. Obesity, 30(3), 738–748. 10.1002/oby.23370

Da Costa, M. (2023). How culture impacts health: The Hispanic narrative. Creative Nursing, 29(3), 273–280. 10.1177/10784535231211695

Emmerich, S. D., Fryar, C. D., Stierman, B., & Ogden, C. L. (2024). Obesity and severe obesity prevalence in adults: United States, August 2021–August 2023 (NCHS Data Brief No. 508). Hyattsville, MD: National Center for Health Statistics. 10.15620/cdc/159281

Endicott, J., Spitzer, R. L., Fleiss, J. L., & Cohen, J. (1976). The Global Assessment Scale: A procedure for measuring overall severity of psychiatric disturbance. Archives of General Psychiatry, 33(6), 766–771. 10.1001/archpsyc.1976.01770060086012

Flórez, K. R., Bell, B. M., Gálvez, A., Hernández, M., Verdaguer, S., & de la Haye, K. (2023). Nosotros mismos nos estamos matando/We are the ones killing ourselves: Unraveling individual and network characteristics associated with negative dietary acculturation among Mexican Americans in New York City. Appetite, 184, 106488. 10.1016/j.appet.2023.106488

George, S., Duran, N., & Norris, K. (2014). A systematic review of barriers and facilitators to minority research participation among African Americans, Latinos, Asian Americans, and Pacific Islanders. American Journal of Public Health, 104(2), e16–e31. 10.2105/AJPH.2013.301706

Ghaddar, S., Brown, C. J., Pagán, J. A., & Díaz, V. (2010). Acculturation and healthy lifestyle habits among Hispanics in United States–Mexico border communities. Revista Panamericana de Salud Pública, 28(3), 190–197. 10.1590/s1020-49892010000900009

Goel, M. S., McCarthy, E. P., Phillips, R. S., & Wee, C. C. (2004). Obesity among US immigrant subgroups by duration of residence. JAMA, 292(23), 2860–2867. 10.1001/jama.292.23.2860

Heredia, N. I., Thrift, A. P., & Balakrishnan, M. (2022). Perceived barriers to weight loss among Hispanic patients with non alcoholic fatty liver disease. Hispanic Health Care International, 20(3), 171–178. 10.1177/15404153211043885

Iruretagoyena, B., Castañeda, C. P., Undurraga, J., Nachar, R., Mena, C., Gallardo, C., … Gonzalez Valderrama, A. (2019). High prevalence of metabolic alterations in Latin American patients at initial stages of psychosis. Early Intervention in Psychiatry, 13(6), 1382–1388. 10.1111/eip.12777

Isasi, C. R., Ayala, G. X., Sotres Alvarez, D., Madanat, H., Penedo, F., Loria, C. M., … Schneiderman, N. (2015). Is acculturation related to obesity in Hispanic/Latino adults? Results from the Hispanic Community Health Study/Study of Latinos. Journal of Obesity, 2015, 186276. 10.1155/2015/186276

Koh, L. M. (2023). Culturally tailoring plant-based nutrition interventions for Hispanic/Latino adults at risk for or with type 2 diabetes: An integrative review. Hispanic Health Care International, 21(2), 89–103. 10.1177/15404153221085696

Li, N., Xue, H., Li, Y., Gao, M., Yu, M., An, C., & Wang, C. (2023). Correlation of obesity and clinical characteristics in drug naive first episode patients with schizophrenia. Clinical Neuropharmacology, 46(5), 186–191. 10.1097/WNF.0000000000000556

Maldonado, L. E., Adair, L. S., Sotres Alvarez, D., Mattei, J., Mossavar Rahmani, Y., Perreira, K. M., … Albrecht, S. S. (2021). Dietary patterns and years living in the United States by Hispanic/Latino heritage in the Hispanic Community Health Study/Study of Latinos (HCHS/SOL). The Journal of Nutrition, 151(9), 2749–2759. 10.1093/jn/nxab165

Morales, L. S., Flores, Y. N., Leng, M., Sportiche, N., Gallegos Carrillo, K., & Salmerón, J. (2014). Risk factors for cardiovascular disease among Mexican American adults in the United States and Mexico: A comparative study. Salud Pública de México, 56(2), 197–205. 10.21149/spm.v56i2.7335

Smith, J., Griffiths, L. A., Band, M., & Horne, D. (2020). Cardiometabolic risk in first episode psychosis patients. Frontiers in Endocrinology, 11, 564240. 10.3389/fendo.2020.564240

Strassnig, M., Kotov, R., Cornaccio, D., Fochtmann, L., Harvey, P. D., & Bromet, E. J. (2017). Twenty-year progression of body mass index in a county-wide cohort of people with schizophrenia and bipolar disorder identified at their first episode of psychosis. Bipolar Disorders, 19(5), 336–343. 10.1111/bdi.12505

Tian, Y., Wang, D., Wei, G., Wang, J., Zhou, H., Xu, H., Dai, Q., Xiu, M., Chen, D., Wang, L., & Zhang, X. Y. (2021). Prevalence of obesity and clinical and metabolic correlates in first episode schizophrenia relative to healthy controls. Psychopharmacology, 238(3), 745–753. 10.1007/s00213-020-05727-1

Vatavuk Serrati, G., Kershaw, K. N., Sotres Alvarez, D., Perreira, K. M., Guadamuz, J. S., Isasi, C. R., Hirsch, J. A., Van Horn, L. V., Daviglus, M. L., & Albrecht, S. S. (2023). Residence in Hispanic/Latino immigrant neighborhoods, away from home food consumption, and diet quality: The Hispanic Community Health Study/Study of Latinos. Journal of the Academy of Nutrition and Dietetics, 123(11), 1596–1605.e2. 10.1016/j.jand.2023.06.283

Ventura, J., Lukoff, D., Nuechterlein, K. H., Liberman, R. P., Green, M. F., & Shaner, A. (1993). Brief Psychiatric Rating Scale (BPRS) expanded version: Scales, anchor points, and administration manual. International Journal of Methods in Psychiatric Research, 3(4), 227–243.

Vintimilla, R., Reyes, M., Johnson, L., Hall, J., & O’Bryant, S. (2020). Factores de riesgo cardiovascular en Estados Unidos y México: comparación de los estudios HABLE y ENASEM. Gaceta Médica de México, 156(1), 17–21. 10.24875/GMM.19005350

