## Supplemental Table 1, 2 and 3. for "Parental Socio-economic Class and Obesity in Hispanic and non-Hispanic Individuals with First Episode Psychosis"

**Supplementary tables**

| Table 1. Baseline Characteristics by Ethnicity Group. | | | | | | | | |
| --- | --- | --- | --- | --- | --- | --- | --- | --- |
| Variable |  | **All (n=145)** |  | **Hispanic (n=24)** |  | **Non-Hispanic (n=121)** |  | ***p*-value** |
| Female sex, n (%) | | 61 (42.06) | | 14 (58.3) | | 47 (38.8) | | 0.123 |
| Age, mean (SD) | | 23.53 (5.29) | | 22.68 (5.32) | | 23.70 (5.29) | | 0.390 |
| Race | |  | |  | |  | | **<0.001*** |
| White, n (%) | | 45 (31.2) | | 13 (56.5) | | 32 (26.4) | |  |
| Black, n (%) | | 64 (44.13) | | 1 (4.3) | | 63(52.1) | |  |
| Asian, n (%) | | 21 (14.48) | | 0 (0.0) | | 21 (17.4) | |  |
| Other, n (%) | | 14 (9.7) | | 9 (39.1) | | 5 (4.1) | | **<0.001*** |
| BPRS, n(SD) | | 46.28 (7.15) | | 46.17 (7.23) | | 46.31 (7.17) | | 0.928 |
| GAS, n(SD) | | 32.76 (5.58) | | 32.17 (4.05) | | 32.90 (5.88) | | 0.563 |
| BMI, mean (SD) | | 23.36 (4.80) | | 25.34 (5.95) | | 22.95 (4.46) | | **0.037*** |
| Primary English, n (%) | | 105 (87.5) | | 15 (75.0) | | 90 (90.0) | | 0.139 |
| Marital Status, n (%) | |  | |  | |  | |  |
| Never Married | | 111 (93.3) | | 18 (90.0) | | 93 (93.9) | | 0.448 |
| Married | | 6 (5.0) | | 1 (5.0) | | 5 (6.1) | |  |
| Divorced | | 2 (1.7) | | 1 (5.0) | | 1 (1.0) | |  |
| Educational Level, n (%) | |  | |  | |  | | 0.386 |
| Less than 7 years | | 35 (29.7) | | 7 (35.0) | | 28 (28.6) | |  |
| Junior high school | | 10 (8.5) | | 0 (0.0) | | 10 (10.2) | |  |
| High school graduate | | 30 (25.4) | | 4 (20.0) | | 26 (26.5) | |  |
| Partial college | | 2 (1.7) | | 0 (0.0) | | 2 (2.0) | |  |
| College graduate | | 29 (24.6) | | 5 (25.0) | | 24 (24.5) | |  |
| Graduate professional school | | 12 (10.2) | | 4 (20.0) | | 8 (8.2) | |  |
| Occupation, n (%) | |  | |  | |  | | 0.640 |
| Semi/Skilled Manual | | 16 (13.7) | | 3 (15.0) | | 13 (13.4) | |  |
| Sales workers | | 31 (26.5) | | 8 (40.0) | | 23 (23.7) | |  |
| Administrative personnel | | 13 (11.1) | | 1 (5) | | 12 (12.4) | |  |
| Business managers | | 27 (23.1) | | 4 (20.0) | | 23 (23.7) | |  |
| High executives | | 22 (18.8) | | 4 (20.0) | | 18 (18.6) | |  |
| SES, mean (SD) | | 44.70 (12.23) | | 46.35 (9.50) | | 44.36 (12.74) | | 0.510 |
| SES, class (%) | |  | |  | |  | | 0.766 |
| Very high class - V | | 21 (17.9) | | 4 (20.0) | | 17 (17.5) | |  |
| High class - IV | | 25 (21.4) | | 4 (20.0) | | 21 (21.6) | |  |
| Middle class - III | | 55 (47.0) | | 11 (55.0) | | 44 (45.4) | |  |
| Low class - II | | 13 (11.1) | | 1 (5.0) | | 12 (12.4) | |  |
| Very low class - I | | 3 (2.6) | | 0 (0.0) | | 3 (3.1) | |  |
| Legend: n=Number of Subjects; %=Percentage; SD=Standard Deviation; BMI=Body Mass Index; BPRS=Brief Psychotic Rating Scale; SES=Socioeconomic Status. | | | | | | | | |

| Table 2. Baseline Characteristics by Ethnicity group – Parents. | | | |
| --- | --- | --- | --- |
| Variable | **Hispanics (n=24)** | **Non-Hispanics (n=121)** | **p-value** |
| BMI, mean (SD) | 25.34 (5.95) | 22.95 (4.46) | 0.037 |
| Education father (%) |  |  | **0.055** |
| Graduate and Professional School | 0 (0.0) | 14 (14.0) |  |
| College Graduate | 2 (10.0) | 19 (19.0) |  |
| Partial College | 1 (5.0) | 16 (16.0) |  |
| High School Graduate | 7 (35.0) | 26 (26.0) |  |
| Partial High School | 3 (15.0) | 3 (3.0) |  |
| Junior High School | 0 (0.0) | 2 (2.0) |  |
| Less than 7 years | 7 (35.0) | 20 (20.0) |  |
| Education mother (%) |  |  | **0.270** |
| Graduate and Professional School | 0 (0.0) | 14 (14.0) |  |
| College Graduate | 3 (15.0) | 28 (28.0) |  |
| Partial College | 4 (20.0) | 16 (16.0) |  |
| High School Graduate | 9 (45.0) | 24 (24.0) |  |
| Partial High School | 0 (0.0) | 2 (2.0) |  |
| Junior High School | 0 (0.0) | 1 (1.0) |  |
| Less than 7 years | 4 (20.0) | 15 (15.0) |  |
| Occupation father (%) |  |  | **0.037** |
| High executives | 0 (0.0) | 8 (9.4) |  |
| Business managers | 0 (0.0) | 25 (29.4) |  |
| Administrative personnel | 4 (23.5) | 10 (11.8) |  |
| Sales workers | 1 (5.9) | 9 (10.6) |  |
| Skilled manual employees | 5 (29.4) | 14 (16.5) |  |
| Semi/Skilled employees | 5 (29.4) | 17 (20.0) |  |
| Unskilled employees | 2 (11.8) | 2 (2.4) |  |
| Occupation mother (%) |  |  | **<0.001** |
| High executives | 1 (5.3) | 0 (0.0) |  |
| Business managers | 1 (5.3) | 34 (37.8) |  |
| Administrative personnel | 6 (31.6) | 20 (22.2) |  |
| Sales workers | 1 (5.3) | 12 (13.3) |  |
| Skilled manual employees | 3 (15.8) | 9 (10.0) |  |
| Semi/Skilled employees | 4 (21.1) | 15 (16.7) |  |
| Unskilled employees | 3 (15.8) | 0 (0.0) |  |
| SES parents, mean (SD) | 31.56 (12.63) | 43.24 (13.59) | **0.002** |
| SES parents class (%) |  |  | **0.028** |
| Very high class - V | 1 (1.4) | 15 (20.3) |  |
| High class - IV | 1 (6.7) | 17 (23.0) |  |
| Middle class - III | 1 (6.7) | 29 (39.2) |  |
| Low class - II | 5 (33.3) | 12 (16.2) |  |
| Very low class - I | 8 (53.3) | 1 (1.4) |  |
| Legend: n=Number of Subjects; %=Percentage; SD=Standard Deviation; BMI=Body Mass Index; BPRS=Brief Psychotic Rating Scale; SES=Socioeconomic Status. | | | |

| Table 3. Stratified by BMI ≥ 25 and < 25 | | | |
| --- | --- | --- | --- |
| Variable | **BMI ≥ 25 (n=36)** | **BMI < 25 (n=87)** | **p-value** |
| Age, mean (SD) | 25.96 (6.82) | 22.85 (4.52) | **0.004*** |
| Ethnicity, n (%) |  |  | **0.005*** |
| Hispanic | 12 (33.3) | 9 (10.3) |  |
| Non-Hispanic | 24 (66.7) | 78 (89.7) |  |
| Female sex, n (%) | 15 (41.7) | 36 (41.4) | 1.000 |
| Race, n (%) |  |  | 0.527 |
| Asian or Pacific Islander | 3 (8.6) | 13 (14.9) |  |
| Black | 14 (40.0) | 41 (47.1) |  |
| Other | 5 (14.3) | 8 (9.2) |  |
| White | 13 (37.1) | 25 (28.7) |  |
| BPRS, mean (SD) | 44.47 (6.31) | 46.58 (7.15) | 0.129 |
| GAS, mean (SD) | 32.34 (5.40) | 32.95 (5.65) | 0.588 |
| Marital status, n (%) |  |  | 0.101 |
| Single | 29 (85.3) | 77 (96.2) |  |
| Married | 4 (11.8) | 2 (2.5) |  |
| Divorced | 1 (2.9) | 1 (1.2) |  |
| Occupation, n (%) |  |  | 0.277 |
| Administrative personnel | 5 (15.2) | 7 (8.9) |  |
| Business managers | 9 (27.3) | 18 (22.8) |  |
| High executives | 9 (27.3) | 11 (13.9) |  |
| Sales workers | 5 (15.2) | 25 (31.6) |  |
| Semi/Skilled employees | 1 (3.0) | 3 (3.8) |  |
| Skilled manual employees | 4 (12.1) | 11 (13.9) |  |
| Unskilled employees | 0 (0.0) | 4 (5.1) |  |
| Unemployed | 0 (0.0) | 4 (5.1) |  |
| Education, n (%) |  |  | 0.111 |
| College Graduate | 6 (18.2) | 22 (27.5) |  |
| Graduate and Professional School | 4 (12.1) | 8 (10.0) |  |
| High School Graduate | 4 (12.1) | 25 (31.2) |  |
| Junior High School | 3 (9.1) | 6 (7.5) |  |
| Less than 7 years | 15 (45.5) | 18 (22.5) |  |
| Partial college | 1 (3.0) | 1 (1.2) |  |
| English Primary Language, n (%) | 28 (82.4) | 72 (90.0) | 0.409 |
| SES, mean (SD) | 46.76 (11.09) | 43.77 (12.59) | 0.239 |
| SES Class, n (%) |  |  | 0.550 |
| Class I | 0 (0.0) | 3 (3.8) |  |
| Class II | 3 (9.1) | 10 (12.7) |  |
| Class III | 14 (42.4) | 38 (48.1) |  |
| Class IV | 10 (30.3) | 15 (19.0) |  |
| Class V | 6 (18.2) | 13 (16.5) |  |
| Education father, n (%) |  |  | 0.141 |
| Graduate and Professional School | 5 (14.7) | 8 (10.0) |  |
| College Graduate | 4 (11.8) | 16 (20.0) |  |
| Partial College | 2 (5.9) | 14 (17.5) |  |
| High School Graduate | 10 (29.4) | 22 (27.5) |  |
| Junior High School | 3 (8.8) | 3 (3.8) |  |
| Less than 7 years | 8 (23.5) | 17 (21.2) |  |
| Education mother, n (%) |  |  | 0.191 |
| Graduate and Professional School | 4 (12.1) | 10 (12.3) |  |
| College Graduate | 4 (12.1) | 26 (32.1) |  |
| Partial College | 5 (15.2) | 15 (18.5) |  |
| High School Graduate | 11 (33.3) | 19 (23.5) |  |
| Partial High School | 1 (3.0) | 1 (1.2) |  |
| Junior High School | 1 (3.0) | 0 (0.0) |  |
| Less than 7 years | 7 (21.2) | 10 (12.3) |  |
| Occupation father, n (%) |  |  | 0.920 |
| High executives | 2 (6.9) | 6 (8.8) |  |
| Business managers | 6 (20.7) | 17 (25.0) |  |
| Administrative personnel | 4 (13.8) | 10 (14.7) |  |
| Sales workers | 2 (6.9) | 7 (10.3) |  |
| Skilled manual employees | 7 (24.1) | 11 (16.2) |  |
| Semi/Skilled employees | 6 (20.7) | 15 (22.1) |  |
| Unskilled employees | 2 (6.9) | 2 (2.9) |  |
| Occupation mother, n (%) |  |  | **0.001*** |
| High executives | 1 (3.1) | 0 (0.0) |  |
| Business managers | 3 (9.4) | 29 (40.3) |  |
| Administrative personnel | 5 (15.6) | 21 (29.2) |  |
| Sales workers | 6 (18.8) | 7 (9.7) |  |
| Skilled manual employees | 4 (12.5) | 7 (9.7) |  |
| Semi/Skilled employees | 11 (34.4) | 8 (11.1) |  |
| Unskilled employees | 2 (6.2) | 0 (0.0) |  |
| SES parents, mean (SD) | 36.08 (14.62) | 43.58 (12.84) | **0.018*** |
| SES parents Class, n (%) |  |  | NaN |
| Class I | 0 (0.0) | 0 (0.0) |  |
| Class II | 7 (30.4) | 12 (19.4) |  |
| Class III | 10 (43.5) | 24 (38.7) |  |
| Class IV | 4 (17.4) | 13 (21.0) |  |
| Class V | 2 (8.7) | 13 (21.0) |  |

**Legend**: n=Number of Subjects; %=Percentage; SD=Standard Deviation; BMI=Body Mass Index; BPRS=Brief Psychiatric Rating Scale; GAS=Global Assessment Scale; SES=Socioeconomic Status; THC=Tetrahydrocannabinol.
*Statistically significant at p<0.05
